# Clinical, social, and economic impacts of colorectal cancer screening with the multi-target stool-DNA test: 10-year experience - a simulated study

**DOI:** 10.1101/2024.08.07.24311643

**Authors:** Chris Estes, Mohammad Dehghani, A. Burak Ozbay, Vahab Vahdat, Paul J. Limburg, Durado Brooks

**Author notes:** **Corresponding author:** Durado Brooks.

## Abstract

**Introduction:** In the United States, colorectal cancer (CRC) remains a substantial public health challenge, with approximately 150,000 new cases diagnosed annually. Guidelines from organizations such as the United States Preventive Services Task Force (USPSTF) recommend several screening strategies, including endoscopic, radiologic, and stool-based options such as the multi-target stool DNA (mt-sDNA) test. In this analysis, we analyzed the estimated clinical, social, and economic impacts of mt-sDNA screening over the inaugural 10-year period for test availability.

**Methods:** To assess the effectiveness of CRC screenings with the mt-sDNA test, published rates of advanced precancerous lesions (APL) and cancer prevalence by stage, as well as the transition rate of APL to CRC for the average-risk population in the US were used for a simulated population. The mt-sDNA test’s sensitivity and specificity for APL and CRC were derived from previously published data. To assess the economic impact of screening with mt-sDNA compared to colonoscopy, we utilized data from literature regarding the time and resources required to prepare and complete each test. Furthermore, the costs of treatment according to the stage of colorectal cancer are considered, to show the value of CRC prevention and early detection.

**Results:** Our analysis indicates that mt-sDNA screening detected an estimated 98,000 cases of CRC and 525,000 individuals were found to have APLs, precursors to CRC. When using 10-year survival rate from CRC, it is estimated that the mt-sDNA test led to more than 34,000 patients surviving due to earlier intervention compared to no screening. Furthermore, the mt-sDNA test demonstrated approximately $22.3 billion cost savings compared to no screening, including an estimated $9.7 billion in cancer treatment costs through early CRC detection and an additional $12.6 billion resulting from cancer prevention through APL detection and management.

**Conclusion:** Clinical availability, adoption, and growth of stool-based CRC screening have significantly increased overall screening rates in the US. It is estimated that mt-sDNA utilization will continue to grow, providing a home-based CRC screening solution for millions of screen-eligible US adults over the next decade and beyond.

## INTRODUCTION

In the United States, colorectal cancer (CRC) remains a substantial public health challenge, with approximately 150,000 new cases diagnosed annually and over 50,000 deaths attributed to the disease each year.^1^ However, overall CRC incidence and mortality rates have been gradually declining over the past few decades,^1^ largely due to increased awareness, effective screening options, and advances in treatment modalities. Nevertheless, CRC is the nation’s second-leading cause of cancer-related death and affects people of all races, genders, and ethnicities.

CRC typically develops over a period of 10-20 years through a multistep process that includes identifiable intermediate precancerous lesions (i.e., adenomatous polyps or sessile serrated lesions), providing ample opportunity for preventing cancer or detecting the disease at early stages with broad-scale screening programs. Early detection of CRC can change lives and is the most important predictor of CRC survival.^2^ Guidelines from organizations such as the United States Preventive Services Task Force (USPSTF) recommend several screening strategies for average-risk individuals, beginning at age 45 years,^3^ including endoscopic, radiologic, and stool-based options such as the multi-target stool DNA (mt-sDNA) test.

Performance characteristics for the mt-sDNA (Cologuard^®^) test were established in the multicenter, nearly 10,000-participant DeeP-C study in 2011, which was subsequently published in the *New England Journal of Medicine*.^4^ After completion of this large prospective clinical validation study, the mt-sDNA test received both FDA approval and CMS coverage in 2014, through the first-ever parallel review by these two agencies. The DeeP-C study showed that the mt-sDNA test detects 92.3% of colorectal cancers and 42.4% of advanced precancerous lesions (APL), with an estimated specificity of 86.6% (all non-advanced adenomas, non-neoplastic findings, and negative results on colonoscopy)^4,5^ by analyzing stool for altered DNA and hemoglobin. Since its clinical availability in August 2014, the mt-sDNA test has contributed substantially to increased participation in CRC screening in the US,^6,7^ with the reported screening rate for Americans aged 50-75 years increasing from 59% in 2013 to 72% in 2021.^8^

## METHODS

mt-sDNA test is an FDA-approved, noninvasive stool-based screening test for adults 45 and older who are at average risk for colorectal cancer.^5^ The mt-sDNA test can be delivered to and picked up from patients’ homes, with no pre-test bowel preparation, no time off from work, and no changes to diet or medication required.^5^ Nationwide, about 95% of all mt-sDNA patients have no out-of-pocket costs for screening.^9^ Within a span of 10 years, the mt-sDNA test has been used more than 16 million times, the equivalent of one mt-sDNA test completed every 20 seconds. In this analysis, we analyze and report the estimated clinical, social, and economic impacts of mt-sDNA screening to date. To simulate the effectiveness of CRC screenings with the mt-sDNA test, published rates of APL and cancer prevalence by stage, as well as the transition rate of APL to CRC for the average-risk population in the US were used (**Table 1**). Other inputs such as mt-sDNA and colonoscopy sensitivity and specificity for APL and CRC were derived from previously published data.^4^

**Table 1.** Input values: clinical, social, and economic impact of mt-sDNA screening.

|  | Measure | Value | Source |
| --- | --- | --- | --- |
| Disease burden and natural history | Advanced precancerous lesion (APL) prevalence | 7.6% | Imperiale 2014 <sup>4</sup> |
|  | CRC prevalence | 0.65% | Imperiale 2014 <sup>4</sup> |
|  | Annual transition of APL to CRC | 8% of APL transition to CRC at 10 years | Stryker 1987 <sup>21</sup> |
|  | Distribution of detected symptomatic CRC by stage | 37% (localized); 40% (regional); 23% (distant) | SEER 1975-1999 <sup>22</sup> |
|  | 10-year survival of CRC by stage | 84.7% (localized); 62.6% (regional); 9.4% (distant) | SEER 2000-2020 <sup>22</sup> |
| Screening performance and impact of screening | mt-sDNA sensitivity | APL (42.4%); CRC: 89.7% (Stage I); 100.0% (Stage II); 90.0% (Stage III); 75.0% (Stage IV) | Imperiale 2014 <sup>4</sup> |
|  | Follow-up colonoscopy sensitivity* | APL (94.0%)**; CRC (95.0%) | Knudsen 2021 <sup>23</sup> |
|  | Distribution of mt-sDNA detected CRC by stage | 79.7% (localized); 15.3% (regional); 5.1% (distant) | Imperiale 2014 <sup>4</sup> |
| | CRC cost by stage | \$169,521 (localized); \$361,917 (regional); \$481,708 (distant) | Fitch 2015 <sup>13</sup> |
| | Average hourly wage | \$34.57 | Bureau of Labor Statistics 2024 <sup>12</sup> |
|  | Patient hours per screen | 1 hour (stool test); 36.22 total hours (colonoscopy); 8 work hours (colonoscopy) | Knudsen 2021 <sup>23</sup>; van der Steen 2015 <sup>24</sup> |
|  | Hours averted per mt-sDNA test kit | 0.13 hours (scheduling); 0.22 hours (nursing); 0.07 hours (provider) | Horejsi 2024 <sup>25</sup> |
|  | Touchdowns per NFL game | 2.36 touchdowns per game | StatMuse 2024 <sup>10</sup> |
|  | Smartphone photos taken per day | 6 photos per day | iHeart 2024 <sup>11</sup> |

\*Assumed 100% follow-up colonoscopy adherence
\*\*Based on 10 lesions <6mm (75.0%); 56 lesions 6-10mm (85.0%); 691 lesions >10mm (95.0%)<sup>4,23</sup>

## RESULTS

Our analysis indicates that mt-sDNA screening detected an estimated 98,000 cases of colorectal cancer (CRC), with nearly 77,00 patients diagnosed at a localized (stage I or II) stage, offering a higher chance of potentially curative treatment (**Figure 1A**).^9^ Moreover, more than 42,000 CRC patients identified by mt-sDNA screening likely avoided chemotherapy or radiation treatment as a result of early-stage detection. Additionally, around 525,000 individuals were found to have APLs, precursors to CRC, for a combined projection of more than 623,000 patients with CRC and/or APLs detected. Our analyses estimate that the mt-sDNA test may have prevented CRC in more than 39,000 individuals through APL detection (and removal at follow up colonoscopy) over the inaugural 10-year period for test availability (**Figure 1A**). Finally, a negative mt-sDNA test result was delivered nearly 14 million times, offering a simple, safe, and effective way for many individuals to obtain reassurance from this noninvasive screening strategy and take charge of their health.

**Figure 1.**
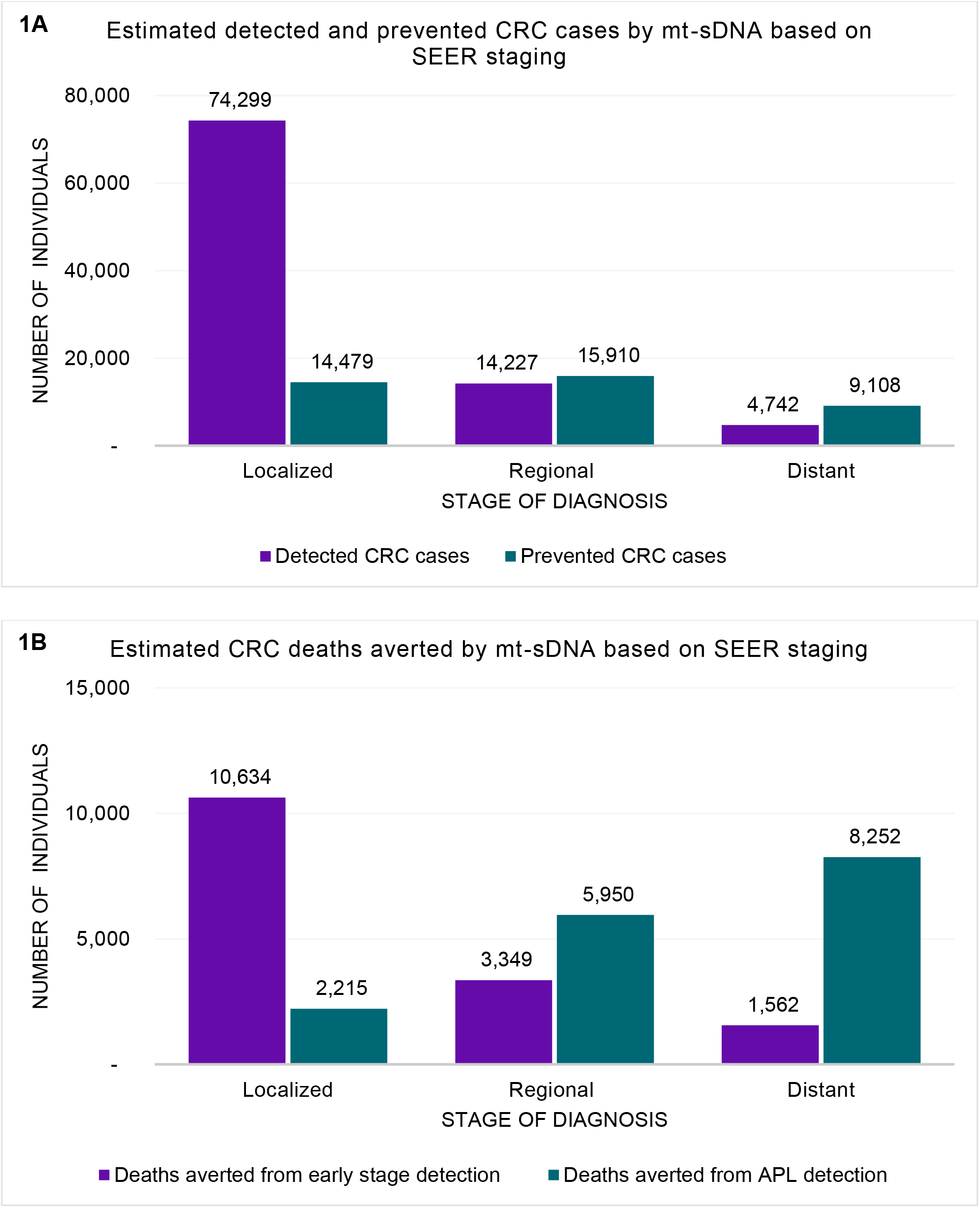
Estimated clinical benefit of screening with mt-sDNA in the US as compared to no screening

When using 10-year survival rate by stage of CRC, it is estimated that the mt-sDNA test led to more than 34,000 patients surviving due to earlier intervention compared to no screening (**Figure 1B**). This finding underscores the significant impact of mt-sDNA screening in improving patient outcomes. For illustration, the additional years gained can be translated to more than 340,000 chances to celebrate a survivor’s birthday, 171,000 more opening or closing ceremony watch parties, 13.7 million more touchdown celebrations,^10^ 205,000 more smartphone photos taken on National Photo Day,^11^ and nearly 125 million more chances to experience a sunrise.

Although colonoscopy has been historically used for the detection of CRC and APL in the US, it is associated with relatively higher costs and associated risks, low screening adherence, and capacity constraints, necessitating the introduction and adoption of noninvasive screening options. To assess the economic impact of screening with the mt-sDNA test compared to colonoscopy, we utilized data from literature regarding the time and resources required to prepare and complete each test (**Table 1**). Taking into account only working hours and an hourly average wage of $34.57 in the US,^12^ the mt-sDNA test was found to save one working day per person screened compared to colonoscopy, resulting in a total estimated wage savings of $4.5 billion over 10 years.

Furthermore, when considering the costs of treatment according to the stage of colorectal cancer,^13^ the mt-sDNA test demonstrated approximately $22.3 billion cost savings compared to no screening, including an estimated $9.7 billion in cancer treatment costs through early CRC detection and an additional $12.6 billion resulting from cancer prevention through APL detection and management. These findings demonstrate the substantial economic benefits of implementing mt-sDNA screening, both in terms of reduced productivity loss and decreased healthcare expenditures associated with treating colorectal cancer.

Finally, the mt-sDNA test reduced hours spent by clinical staff, with more than 16 million screenings resulting in 2.2 million fewer scheduling hours, 3.5 million fewer nursing hours, and 1.1 million fewer provider hours. These reductions have allowed staff to focus limited colonoscopy resources on high-risk and symptomatic individuals.

## DISCUSSION

Clinical availability, adoption, and growth of stool-based CRC screening have significantly increased overall screening rates in the US, with the characteristics and contributions of the mt-sDNA test described in nearly 100 peer-reviewed publications to date.^14^ Notable examples of clinically-relevant research include the reported observation that the mt-sDNA test accounted for 77% of the improvements in CRC screening between 2018 and 2021.^7^ In addition, reported age-adjusted CRC mortality has declined by more than 10% during 2014 to 2022.^15^ The limited capacity for hospital- or clinic-based screening evaluations such as colonoscopy suggests that prioritizing appointments for follow-up as indicated after a positive stool test could provide greater public health benefit.^16^ Research indicates that the colonoscopy backlog in the US could extend up to eight years with approximately 60 million average-risk individuals eligible for screening or re-screening in the US^17,18^; however, this backlog can be much more effectively managed with initial non-invasive screening. Additionally, previous studies have shown that follow-up colonoscopies after positive stool-based tests are not only cost-saving,^19^ but also three times more effective and beneficial than screening colonoscopies.^16^ Because financial barriers were a significant contributor to suboptimal follow-up colonoscopy completion rates in the US,^20^ federal regulations required commercial insurers and Medicare to eliminate out-of-pocket costs for follow-up colonoscopy beginning in 2023. These initiatives alongside the ease, effectiveness, and personalized navigation programs that have been offered by the mt-sDNA test have led to increased utilization in recent years. It is estimated that mt-sDNA utilization will continue to grow, providing a home-based CRC screening solution for millions of screen-eligible US adults over the next decade and beyond.

## Data Availability

All data produced in the present study are available upon reasonable request to the authors

## REFERENCES

1. Siegel RL, Wagle NS, Cercek A, Smith RA, Jemal A. Colorectal cancer statistics, 2023. CA: A Cancer Journal for Clinicians. 2023;73(3):233–254.

2. National Cancer Institute. Cancer Stat Facts: Colorectal Cancer. 2024; https://seer.cancer.gov/statfacts/html/colorect.html. Accessed July 30, 2024.

3. Lin JS, Perdue LA, Henrikson NB, Bean SI, Blasi PR. U.S. Preventive Services Task Force Evidence Syntheses, formerly Systematic Evidence Reviews. In: Screening for Colorectal Cancer: An Evidence Update for the U.S. Preventive Services Task Force. Rockville (MD): Agency for Healthcare Research and Quality (US); 2021.

4. Imperiale TF, Ransohoff DF, Itzkowitz SH, et al. Multitarget stool DNA testing for colorectal-cancer screening. N Engl J Med. 2014;370(14):1287–1297.

5. Exact Sciences Corporation. Cologuard Clinician Brochure. Madison, WI.

6. Cheney C, Parish A, Niedzwiecki D, et al. Colorectal cancer screening uptake and adherence by modality at a large tertiary care center in the United States: a retrospective analysis. Curr Med Res Opin. 2024;40(3):431–439.

7. Ebner DW, Finney Rutten LJ, Miller-Wilson LA, et al. Trends in colorectal cancer screening from the National Health Interview Survey (NHIS): analysis of the impact of different modalities on overall screening rates. Cancer Prev Res (Phila). 2024.

8. National Cancer Institute. Cancer Trends Progress Report - Colorectal Cancer Screening. 2024; https://progressreport.cancer.gov/detection/colorectal_cancer. Accessed July 30, 2024.

9. Exact Sciences Corporation. Internal Data on File. Madison, WI.

10. StatMuse. NFL League Average Touchdowns per Game. 2024; https://www.statmuse.com/nfl/ask/nfl-league-average-touchdowns-per-game.

11. iHeart. SURVEY SAYS: Average American Takes 6 Pics a Day. 2023; https://www.iheart.com/content/2023-09-29-survey-says-average-american-takes-6-pics-a-day/.

12. U.S. Bureau of Labor Statistics. Table B-3. Average hourly and weekly earnings of all employees on private nonfarm payrolls by industry sector, seasonally adjusted. 2024; https://www.bls.gov/news.release/empsit.t19.htm. Accessed May 8, 2024.

13. Fitch K, Pyenson B, Blumen H, Weisman T, Small A. The value of colonoscopic colorectal cancer screening of adults aged 50 to 64. Am J Manag Care. 2015;21(7):e430–438.

14. Pubmed - National Library of Medicine. Cologuard-Related Research since 2014. 2024; https://pubmed.ncbi.nlm.nih.gov/collections/64158012/.

15. National Cancer Institute. SEER*Explorer: An interactive website for SEER cancer statistics - Colon and Rectum Recent Trends in U.S. Age-Adjusted Mortality Rates, 2000-2022. https://seer.cancer.gov/statistics-network/explorer/application.html. Accessed July 30, 2024.

16. Fendrick AM, Borah BJ, Ozbay AB, Saoud L, Limburg PJ. Life-years gained resulting from screening colonoscopy compared with follow-up colonoscopy after a positive stool-based colorectal screening test. Preventive Medicine Reports. 2022;26:101701.

17. Fendrick AM, Ebner D, Kisiel JB, et al. Sa1144 ELIMINATING THE COLONOSCOPY BACKLOG WITH STOOL-BASED COLORECTAL CANCER SCREENING OPTIONS. Gastroenterology. 2024;166(5):S-358-S-359.

18. Ebner DW, Kisiel JB, Fendrick AM, et al. Estimated Average-Risk Colorectal Cancer Screening– Eligible Population in the US. JAMA Network Open. 2024;7(3):e245537–e245537.

19. Fendrick AM, Kisiel JB, Brooks D, et al. A call to action to increase uptake of follow-up colonoscopy after initial positive stool-based colorectal cancer screening. Population health management. 2023;26(6):448–450.

20. Fendrick AM, Princic N, Miller-Wilson LA, Wilson K, Limburg P. Out-of-Pocket Costs for Colonoscopy After Noninvasive Colorectal Cancer Screening Among US Adults With Commercial and Medicare Insurance. JAMA Netw Open. 2021;4(12):e2136798.

21. Stryker SJ, Wolff BG, Culp CE, Libbe SD, Ilstrup DM, MacCarty RL. Natural history of untreated colonic polyps. Gastroenterology. 1987;93(5):1009–1013.

22. National Cancer Institute SEER*Stat software (seer.cancer.gov/seerstat) version 8.4.3 [computer program].

23. Knudsen AB, Rutter CM, Peterse EFP, et al. U.S. Preventive Services Task Force Evidence Syntheses, formerly Systematic Evidence Reviews. In: Colorectal Cancer Screening: An Updated Decision Analysis for the U.S. Preventive Services Task Force. Rockville (MD): Agency for Healthcare Research and Quality (US); 2021.

24. van der Steen A, Knudsen AB, van Hees F, et al. Optimal colorectal cancer screening in states’ low-income, uninsured populations—the case of South Carolina. Health Serv Res. 2015;50(3):768–789.

25. Horejsi A, Roberts C, Walter J, et al. Augmenting Intelligence & Amplifying Health: Proactive Outreach for Enhanced Preventive Screening. Paper presented at: ACHE Congress - American College of Healthcare Executives 2024; Chicago.

